# A polygenic score-based approach to identify gene-drug interactions stratifying breast cancer risk

**DOI:** 10.1101/2021.05.03.21256511

**Authors:** Andrew R. Marderstein, Scott Kulm, Cheng Peng, Rulla Tamimi, Andrew G. Clark, Olivier Elemento

## Abstract

An individual’s genetics can dramatically influence breast cancer (BC) risk. While clinical measures for prevention do exist, non-invasive personalized measures for reducing BC risk are limited. Commonly-used medications are a promising set of modifiable factors, however no previous study has explored whether a range of widely-taken approved drugs modulate BC genetics. In this study, we describe a quantitative framework for exploring the interaction between the genetic susceptibility of BC and medication usage among UK Biobank women. We computed BC polygenic scores (PGS) that summarize BC genetic risk, and find that the PGS explains nearly three-times greater variation in disease risk within corticosteroid users compared to non-users. We map 35 genes significantly interacting with corticosteroid use (*FDR* < 0.1), highlighting the transcription factor NRF2 as a common regulator of gene-corticosteroid interactions in BC. Finally, we discover a novel regulatory variant strongly stratifying BC risk according to corticosteroid use. Within risk allele carriers, 18.2% of women taking corticosteroids developed BC, compared to 5.1% of the non-users (with a *HR* = 3.41 per-allele within corticosteroid users). Overall, this work highlights the clinical relevance of gene-drug interactions in disease risk, and provides a roadmap for repurposing biobanks in drug repositioning and precision medicine.

## Introduction

Breast cancer (BC) is the most commonly diagnosed cancer in women, with over 2 million new cases diagnosed and 600,000 deaths in 2018 worldwide^1^, highlighting a clear need to implement primary prevention strategies. However, preventive measures are limited, with many involving invasive surgical procedures (such as mastectomy) or drugs (such as tamoxifen) with moderate to severe side effects.

Repurposing existing medications that treat other indications provides a unique opportunity to identify novel therapeutic targets for breast cancer risk reduction. However, germline genetic variation is one potential reason for variable drug efficacy and adverse outcomes. For example, vitamin B12 intake is not associated with BC risk among the Women’s Health Study participants^2^ (who are primarily white), but is significantly associated with reduced BC risk among Mexican women^3^ and Canadian *BRCA1* mutation carriers^4^. While these differences may be due to a number of study design and methodologic differences, it may also suggest differences due to genetic background. Given the prevalence of gene-environment interactions^5-7^, medication-associated risk reduction may strongly depend on genetic factors and thus be different between individuals.

Most discovered gene-drug interactions to date involve variation at individual genes^8^ (such as *CYP2C9*- or *VKORC1*-warfarin interactions for anticoagulation^9^, *HMGCR*-statin interactions for cholesterol^10^, or *CYP2D6-* or *SULT1A1*-tamoxifen interactions for breast cancer^11-13^), with the contribution from genome-wide variation largely unexplored. Genome-wide association studies (GWAS) have revealed that the genetic variance for most phenotypes is spread genome-wide at thousands to tens of thousands of loci rather than the most significant single nucleotide polymorphisms (SNPs)^14^. Thus, while individual genes have been shown to strongly influence the response to treatments, the genome-wide polygenic contribution to drug response deserves further exploration. In one analysis of three statin clinical trials, the relative risk reduction of coronary artery disease for those at high polygenic risk was 46%, compared to 26% in all other individuals^15^. Patients with a particular genetic risk profile receiving a drug may experience greater risk protection against disease than other drug users with a different set of genetic alterations. Importantly, genetic activation of a disease pathway may be nullified through a drug’s mechanism of action.

Generally, pharmacogenomic studies—which have successfully identified genes and pathways influencing the body’s response to hundreds of drugs^8^—have been relatively narrow in scope, with specific hypotheses linked to particular mutations or drugs. A systematic search for interactions between commonly-used drugs and genome-wide variants remains unexplored. This could lead to potential drug repurposing or personalized medication prescribing to reduce disease risk within specific subsets of the population (e.g. those who have extremely high polygenic risk^15- 17^ or carry particular risk variants^4,9-11^). Extensive analysis of the pharmacogenomic interactions between a variety of medications and genome-wide data can reveal why some medication users experience adverse clinical outcomes or point to potential therapeutic repurposing opportunities for women with genetic predispositions to breast cancer risk (**Figure 1**).

**Figure 1:**
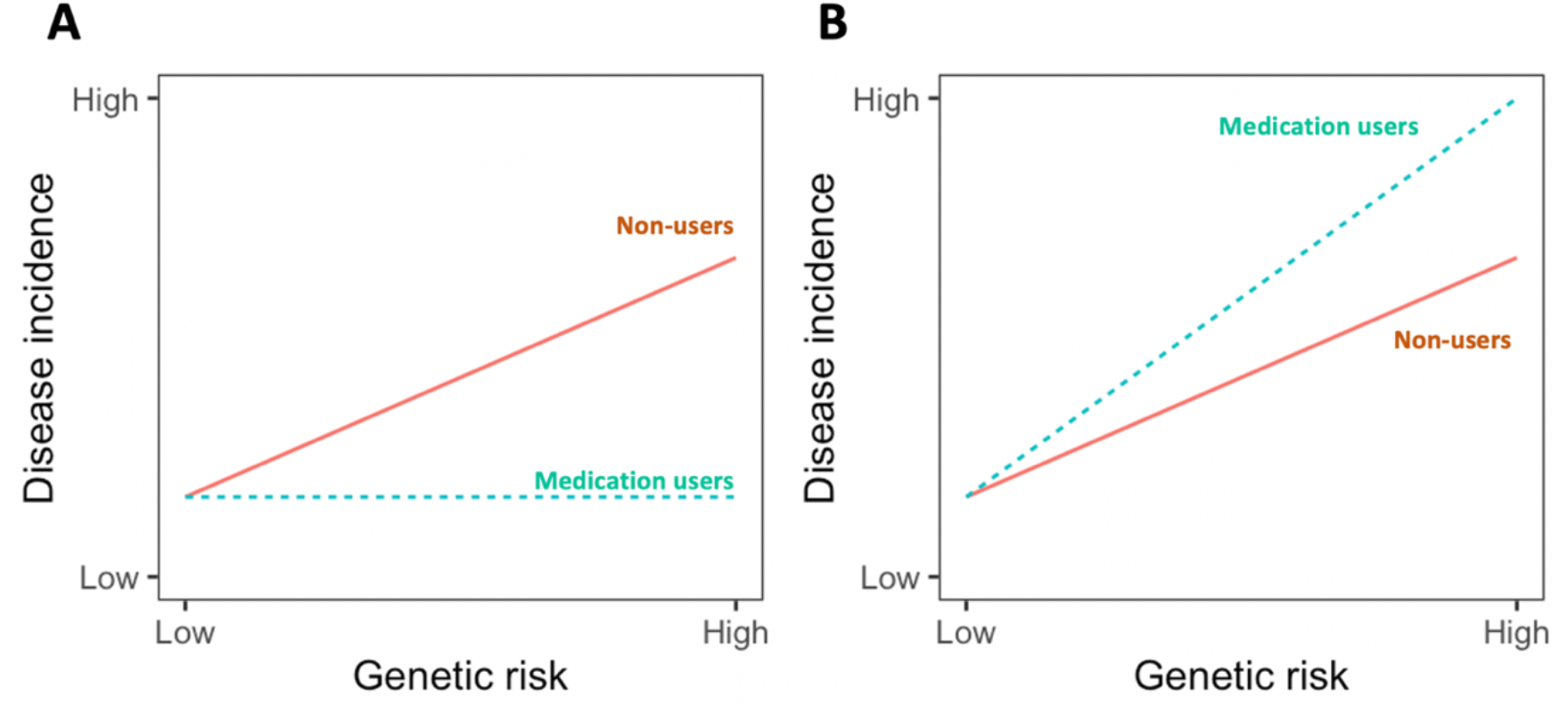
Modulating genetic risk by using modifiable factors. Genetic variants can increase disease risk. Medications are easily modifiable with strong biological effects, with the potential to alter the genetic effects of risk variants. In population studies, this can appear as an interaction, where drug use drastically modifies the association between genetic risk factors and disease rates. It is unclear whether approved and regularly-taken medications can reverse or limit the effects of genetic risk. Figure 1A illustrates an case where the medication ameliorates genetic risk (an antagonistic interaction). However, medication use may instead exacerbate disease risk (Figure 1B; a synergistic interaction). Statins as a means for reducing coronary artery disease risk are one prominent example of an antagonistic interaction^15,18^.

With the emergence of UK Biobank (UKB), a large publicly available dataset that combines electronic health record data with genomic, prescription, and survey questionnaire information from 500,000 individuals, it is possible to study such potential pharmacogenomic interactions at scale. This cohort has already revealed the genetic influences on medication use^19^ and dosage^20,21^, the population prevalence of known pharmacogenomic variation^22^, the incidence of drug side effects^21^, and the genetic and non-genetic characteristics of treatment-resistant depression^23^.

In the present study, we introduce a framework for using UKB to identify interactions between genetic and medication data. We identify corticosteroids as a modulator of polygenic risk in BC, and use a SNP-based gene-set enrichment analysis to highlight potential pathways and mechanisms of this interaction. Finally, we assessed stratification of BC risk in UKB by corticosteroid use and genetic variation. Overall, our results demonstrate the potential of prospective cohorts such as UKB in drug repurposing, risk prediction, and pharmacogenomics by using statistical genetic and epidemiologic approaches (**Figures 2A-B**).

**Figure 2:**
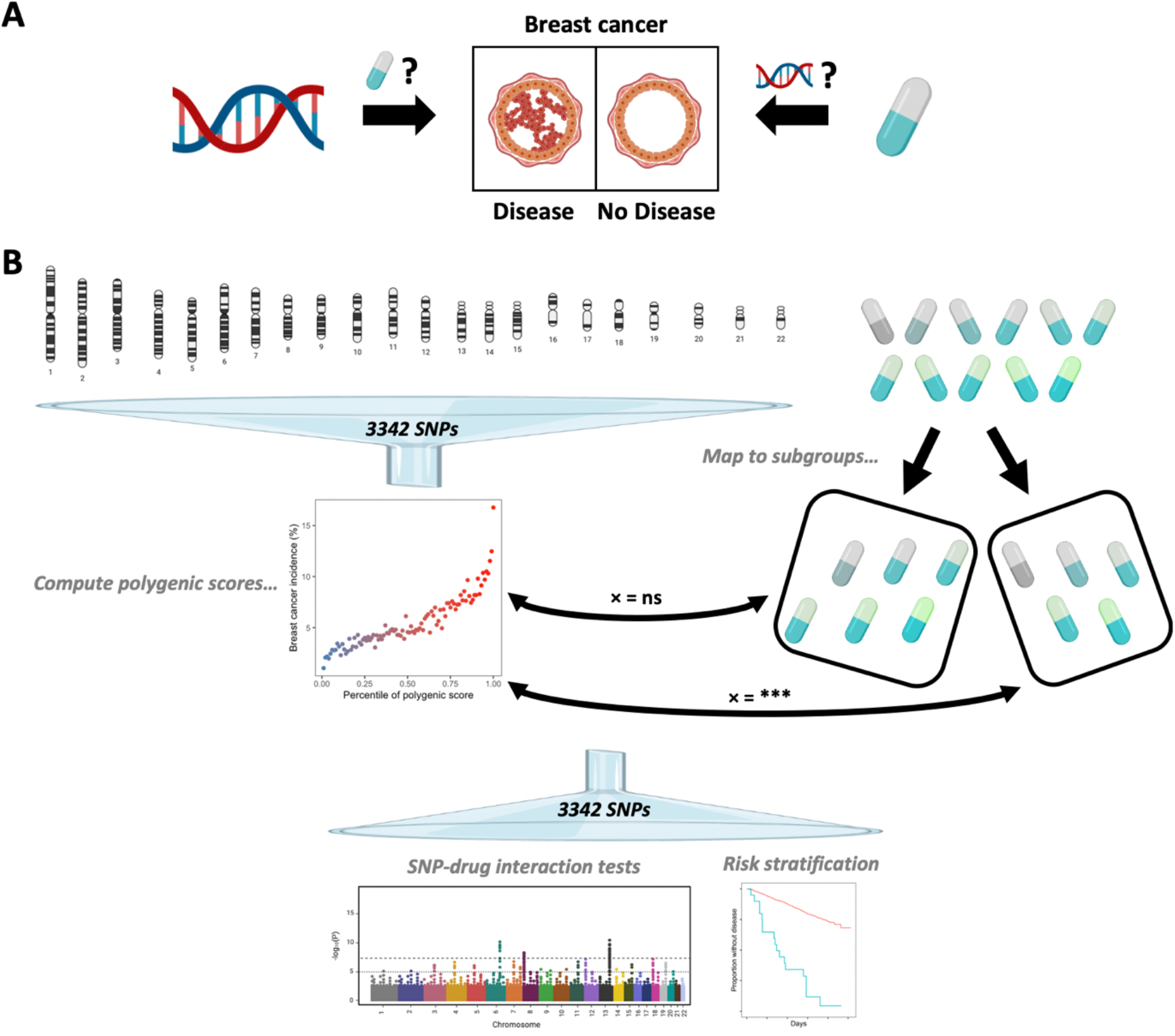
Overview of the study. (A) We searched for drugs that modify the effect of a genetic variant or the effect of polygenic variation on breast cancer risk; or, alternatively, whether there are genetic influences that modify a drug’s risk profile in terms of breast cancer risk. (B) In our study, we summarized effects of breast cancer susceptibility loci across the genome by computing polygenic scores and assembled medications within larger subgroups. Next, we tested for polygenic-drug interactions between polygenic scores and medicines. After identifying significant polygenic-drug interactions, we tested for SNP-drug interactions to identify individual loci driving the statistical signal, infer mechanisms, and stratify breast cancer risk. Manhattan plot is obtained from BioRender and is purely theoretical.

## Results

### Introduction to coordinated interactions through the lens of drug response

In a prior application to epistasis, Sheppard et al.^24^ described how statistical interactions between polygenic scores (PGS) and other factors are driven by SNPs broadly interacting positive or negatively, in a model named a coordinated interaction. Thus, a significant interaction between PGS and drug use (a “polygenic-drug interaction”) with regard to a particular phenotype means that drug use serves to strengthen or dampen—on average—the marginal genetic effects of causal variants. Such a coordinate interaction may be positive or negative. A positive polygenic-drug interaction indicates that SNP effects are on average enhanced (synergistic), while a negative polygenic-drug interaction indicates that SNP effects are on average reduced (antagonistic). In theory, an antagonistic interaction can be clinically relevant for reducing the effects of “poor” (or elevated) genetic risk, while synergistic interactions can help inform why some individuals experience adverse outcomes (**Figure 3**).

**Figure 3:**
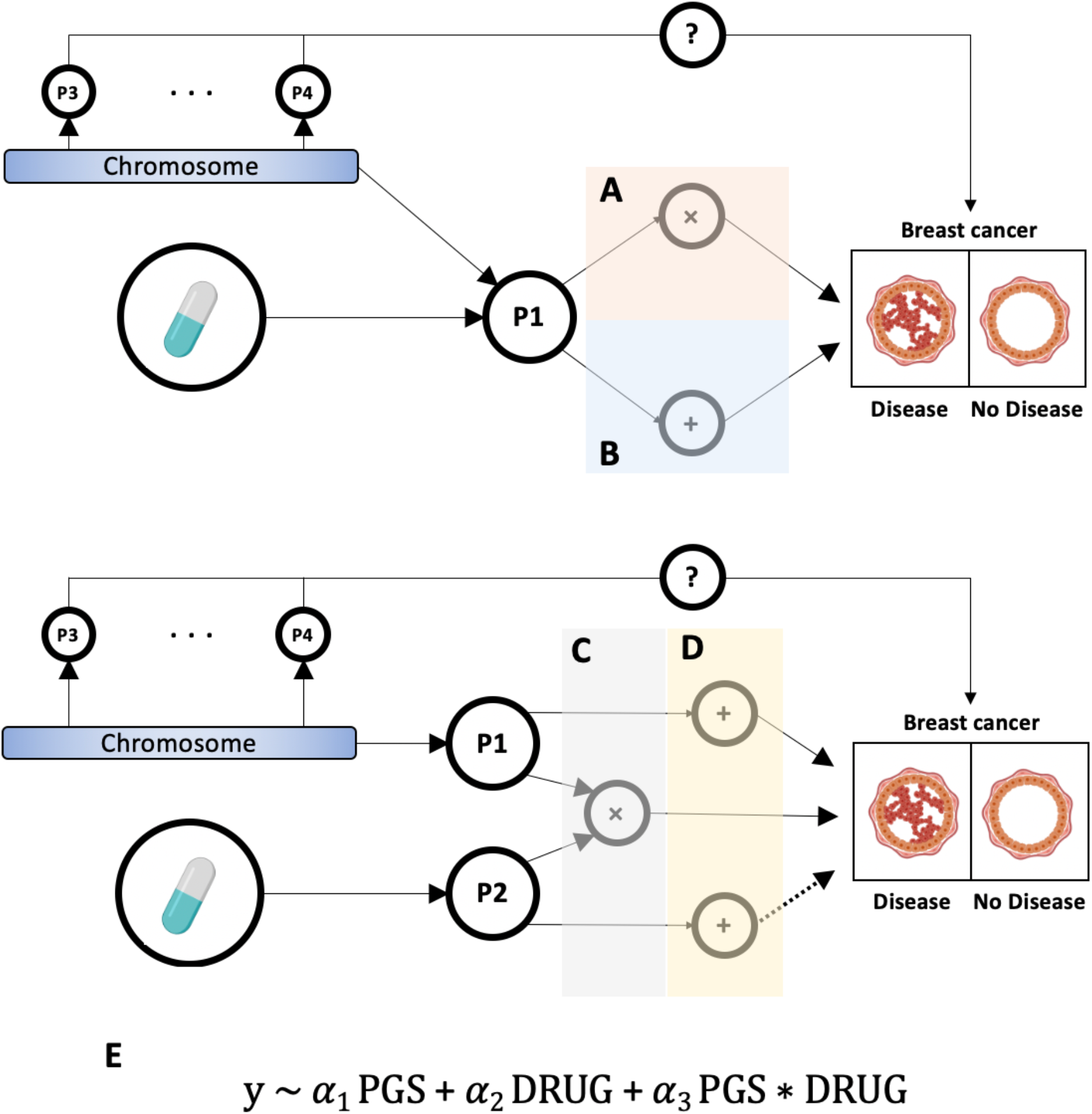
Coordinated polygenic-drug interactions. Polygenic scores include risk variants across the genome, which influence multiple biological pathways (e.g. P1, P3, or P4) contributing to disease risk (either through epistatic or main effects). Drugs may target a similar pathway(s) as a subset of the risk variants (top; P1), or a distinct pathway(s) (bottom; P2). In a scenario where a subset of risk variants and a drug influence the same pathway (top), then there may exist interaction effects (A)—where, for example, the drug may help nullify genetic risk factors (antagonistic interactions), or the drug may drastically magnify the genetic risk factors (synergistic interactions). The effects can also be independent, and thus no interaction effects exist (B). If the drug targets a distinct pathway(s) compared to the genetic risk factors (bottom; P2), then an interaction might be present if the two pathways interact (C). In these cases, protective medication effects (antagonistic interactions) or harmful medication effects (synergistic interactions) may only exist with genetic perturbations in the core disease pathways. However, the drug-targeted pathways may also never interact with the disease pathways affected by common genetic variants, and thus medication use would be either independently associated with disease risk or have no association altogether (D). Any potential interactions are captured by a coordinated polygenic-drug interaction test (E), where the interaction between medication use (DRUG) and genome-wide variation on disease risk (y) is tested for by aggregating directional signal across variants in a polygenic score (PGS).

If the SNP main effects have no correlation with the SNP interaction effects with the drug (“uncoordinated interactions”), then there is no power to detect a significant interaction. We demonstrate this point in the Supplement, where we found across simulations that the estimated interaction effects between PGS and drug use are correlated to the covariance between true main and interaction effects; furthermore, power to detect interactions increases as this correlation increases (Supplementary Notes 1-3; Supplementary Figure 1). Based on this, we can conclude that testing for a polygenic-drug interaction is an effective approach for assessing the relationship between main and interaction effects, and thereby identifying drugs which modulate genetic risk.

### Computing a polygenic score for breast cancer in UK Biobank

Within UKB, we identified a cohort of 212,335 disease-free women who had no self-reported breast cancer or previous diagnosis of breast cancer in their hospital records at the time of baseline assessment (Supplementary Figure 2). Of the 212,335 disease-free women at baseline, 11,730 incident developed incidence BC over the course of follow-up (determined through longitudinal health records).

We use a PGS to summarize BC risk from common genetic variation in the population. Interaction analyses often suffer from low power, and PGS vary between score generation methodologies and summary statistics used. With this in mind, we aimed to create an optimal PGS in terms of bias, accuracy, and robustness. Using the summary statistics from Michailidou et al.^25^ and the PGS weights provided by Mavaddat et al.^26^ in the PGS catalog^27^ (Mavaddat et al. did not report the original GWAS summary statistics), we calculated multiple BC scores for all individuals from both studies. We then selected the optimal score from Michailidou et al. and the optimal score from Mavaddat et al. by including each score separately in a BC risk model and selecting the score with the maximal variation explained (via Nagelgerke-R^2^) (Supplementary Figure 3).

Interestingly, we found that the Michailidou et al. and Mavaddat et al. scores had a weaker correlation than initially expected (*r* = 0.59; **Figure 4A**). For example, only 40.1% of individuals within the top decile of the Michailidou et al. score were within the top decile of the Mavaddat et al. score, and only 25.1% of individuals classified in the top decile of either score were in the top decile of both scores. As an accurate PGS will power the downstream interaction analyses, and owing to bias in the score due to differences in polygenic scoring method, we tested whether an ensemble score between the two would improve PGS accuracy. We merged standardized Michailidou et al. and Mavaddat et al. scores into a combined score by averaging the two, and assessed the variation explained by the three scores.

**Figure 4:**
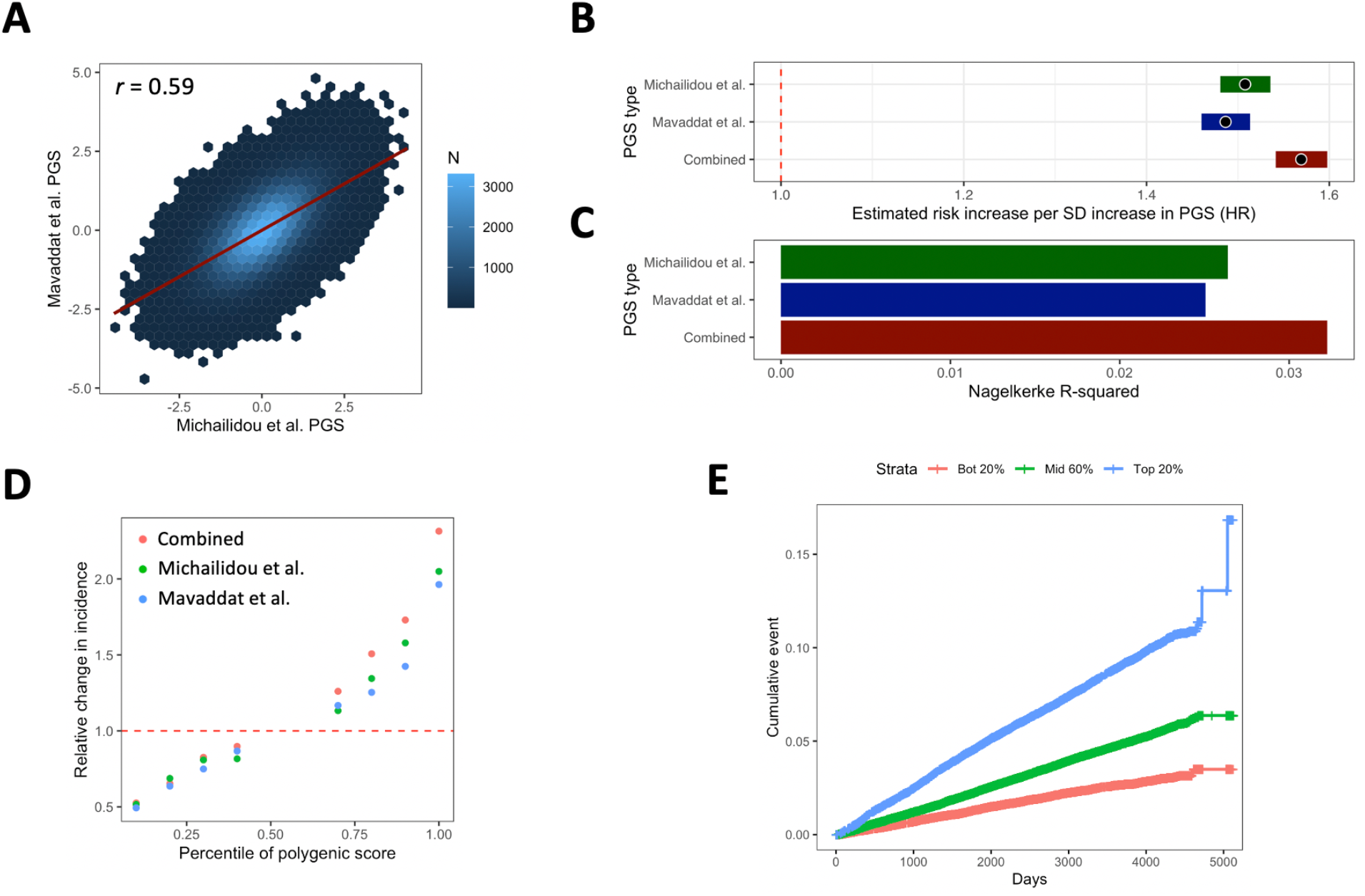
Improving PGS accuracy by combining scores. (A) A density map of the correlation between the optimal Michailidou et al. PGS and the optimal Mavaddat et al. PGS. Correlation is strong, but modest. (B) Hazard ratio estimates with 95% CI errors for the Michailidou et al., Mavaddat et al., and combined scores per standard deviation increase. (C) Nagelkerke R-squared for a model with Michailidou et al., Mavaddat et al., and combined PGS compared to a model without PGS. (D) Breast cancer incidence within deciles relative to the middle 45^th^ to 55^th^ percentile women, for the combined, Michailidou et al., and Mavaddat et al. PGSs. 40-50^th^ and 50-60^th^ deciles are removed. (E) Cumulative rate of breast cancer (y-axis) as a function of days (x-axis), stratified across three groups: top 20% of the PGS, middle 60% of the PGS, and bottom 20% of the PGS.

We found that the combined score was an improved predictor and explained greater variation in BC risk compared to the Michailidou et al. or Mavaddat et al. scores. The estimated risk increase in Cox Proportional-Hazards models was *HR*=1.569 (95% CI: 1.541-1.598) per standard deviation increase in the combined score, compared to a *HR*=1.508 (95% CI: 1.481-1.535) and *HR*=1.486 (95% CI: 1.460-1.513) increase in the Michailidou et al. and Mavaddat et al. scores (**Figure 4B**). Furthermore, the combined score explained 22.2% and 28.7% more variation in BC risk compared to the Michailidou et al. and Mavaddat et al. scores, as determined by Nagelkerke R-squared (**Figure 4C**). Lastly, we found that the incidence of BC for women in the top 10% of the combined score was 2.31-times higher rate than that of women between the 45^th^ and 55^th^ score percentiles (*P* < 10^−200^). In comparison, the incidence in the top 10% of the Michailidou et al. and Mavaddat et al. scores was 2.04- and 1.96-times higher rate than in the 45^th^ to 55^th^ percentile within each score respectively (**Figure 4D)**. Overall, the combined PGS for BC effectively stratified risk and timing of disease onset: 2.8%, 5.1%, and 9.5% of women in the bottom 20%, middle 60%, and top 20% of PGS developed BC (**Figure 4E**). Thus, we next assessed whether medication usage influenced genetic risk of BC in the UKB participants by specifically testing the interaction between PGS and medication use.

### Identifying polygenic-drug interactions associated with breast cancer

At recruitment, UKB individuals reported the medications taken regularly (Supplementary Figure 2), reporting 3,603 unique medications. Using the mapping provided in Wu et al.^19^, the 3,603 medications were mapped to the Anatomical Therapeutic Chemical (ATC) Classification System^28^ and grouped into the Level 4 ATC codes (**Figure 2B**). This left 106 therapeutic subgroups with at least 1,000 users in UKB. However, treatments are not randomly assigned to individuals (such as in a randomized controlled trial). We removed 4 medication groups (A03FA, G02BA, G03CX, G03FA) with strong marginal associations with BC risk (*FDR* < 0.01; see Methods), since it is unclear whether interactions with these medications are more likely to be due to risk factors that have not been accounted for in analysis (such as the drug indication) or the drug itself. The G0*** medications are related to birth control, while A03FA medications (Propulsives) are used for gastrointestinal complications, such as heartburn, nausea, or vomiting.

Next, we sought to detect polygenic score-drug interactions. We assessed the pairwise interaction with 102 medication groupings on future BC diagnosis, by including an interaction term representing the interaction between PGS and a drug within 102 distinct Cox Proportional-Hazards models, along with main effects for score, drug, and various covariates (see Methods). Overall, we found that 14 of 102 medication groups tested have a nominal *P* < 0.05 interaction with PGS on BC risk (13.7%) (**Figure 5A**). After multiple testing correction, we identified 5 significant interactions (*FDR* < 0.1) with the following ATC groups: S01BA, D07XA, D07AA, S02BA, and H02AB. These groups all relate to corticosteroids and are highly correlated due to drugs mapping to multiple ATC annotations, so we used only S01BA for further analysis of corticosteroids (*FDR* = 0.03). The polygenic-drug interaction with corticosteroids is synergistic, where the risk increase per standard deviation increase in the PGS was 38.5% higher in the users (*HR* = 2.16) compared to non-users (*HR* = 1.56). As a result, the PGS explains nearly three-times greater variation in BC risk within corticosteroid users (Nagelkerke-R^2^ = 0.090) compared to non-users (Nagelkerke-R^2^ = 0.032) (**Figure 5B**). This demonstrates that lifestyle factors, such as medication usage, considerably influence the genetic contribution to risk.

**Figure 5:**
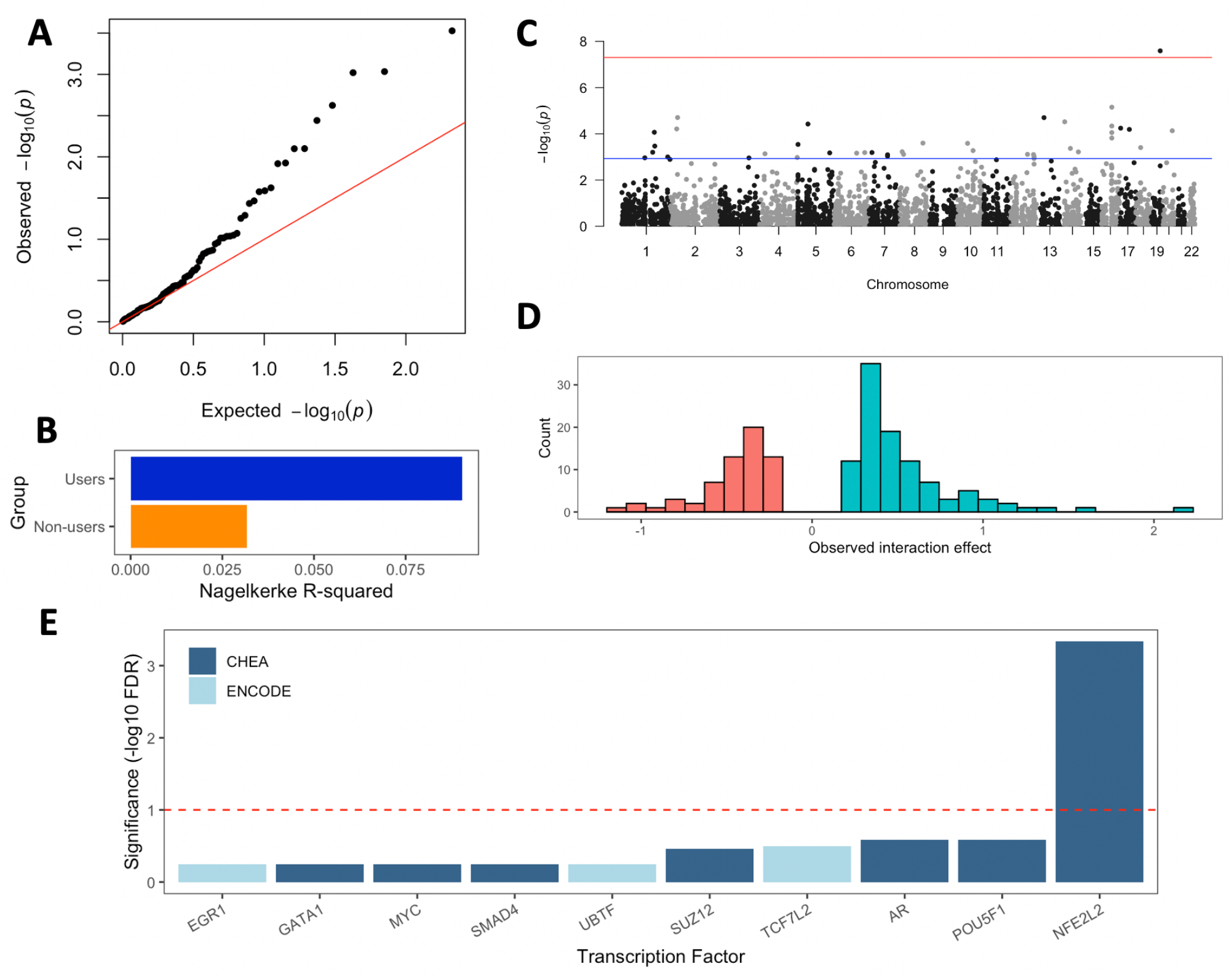
Gene-drug interactions associated with breast cancer risk. (A) Quantile-quantile plot containing 102 interaction tests between polygenic score and 102 medication groups. Each point represents a different drug and the −log10 *P*-value of the interaction. (B) The Nagelkerke R-squared of the PGS within the corticosteroid users versus non-users in UKB. (C) Manhattan plot of the SNP-corticosteroid interaction tests. Each point represents a different SNP and the −log10 *P*-value of the interaction. The blue line represents *FDR* < 0.1, and the red line represents *P* < 5 x 10^−8^. (D) A histogram of the estimated interaction effect for all risk-increasing SNPs with an interaction *P* < 0.05. (E) Gene-set enrichment of interaction genes for common transcription factor regulators. Interaction genes are determined by identifying all SNPs containing an interaction with *FDR* < 0.1, and mapping SNPs to genes. Analysis performed using EnrichR and the ENCODE_and_ChEA_Consensus_TFs_from_ChIP-X” database.

### Polymorphisms interact with corticosteroid use and drive the polygenic signal

The disease mechanisms implicated by PGS are unclear, and so are the biological processes contributing to an interaction with corticosteroids. These scores include genome-wide susceptibility loci often lying in multiple biological pathways. Given that we observe a significant interaction with corticosteroids, we considered whether it would be possible to identify potential subsets of BC genes, pathways, or mechanisms which interact with corticosteroid exposure. Thus, we looked for particular SNPs which may drive the observed polygenic-drug interaction signal, which can provide causal insights into human biology. We took the 3,342 SNPs used to compute the combined PGS and tested for pairwise interactions between a SNP and corticosteroid use within 3,342 distinct interaction models (**Figure 5C**).

Overall, we found many SNPs involved in these interactions. After multiple testing correction, the interaction term for 1.1% of tested SNPs are significant at *FDR* < 0.1 (40 of 3,342 SNPs) (Supplementary Table 1). Statistically, we also found evidence for coordinated interactions on the SNP-level by considering the SNPs with interaction *P*-values below 0.05. Within the set of risk-increasing variants (from a marginal GWAS), we found that synergistic SNP-drug interactions are significantly enriched, while antagonistic SNP-drug interactions are depleted (62.2% versus 37.8%; *P* = 2.2 x 10^−3^) (**Figure 5D**). No significant trend was observed for risk-decreasing variants. It appears that many SNPs are driving the statistical interaction we observe with polygenic risk.

### Functional insights into corticosteroid interactions with breast cancer risk

Next, we aimed to identify potential mechanisms and pathways which interact with corticosteroids and modulate BC risk. We first assigned the 40 SNPs with significant interactions (*FDR* < 0.1) to 35 genes (Supplementary Table 1) by using the multi-omics-based V2G pipeline in Open Targets^29^, and performed a number of gene-set enrichment analyses by using EnrichR^30^ (see Methods).

We first assessed whether the 35 interaction genes overlapped with collected drug-gene signatures in DSigDB, which are based on drug-induced gene expression changes. While there was no significant overlap with gene expression changes from approved agonists of the glucocorticoid receptor (GR) (the molecular target for many corticosteroids), we identified a significant enrichment for mifepristone, an approved GR antagonist, in DSigDB after correcting for all drug-gene signatures in DSigDB^30,31^ (*FDR* = 0.09). Mifepristone (also known as RU-486) blocks progesterone and is currently used for terminating pregnancies; interestingly, its use as a treatment for BC is being explored within clinical trials^32^ based on promising experimental data^33,34^.

Next, we assessed enrichment of gene ontology (GO) terms for the 35-interaction gene set. We found enriched GO terms related to transcriptional processes, such as transcriptional coactivator binding, and the regulation of signal transduction pathways, such as the Wnt signaling pathway previously implicated in cancer development^35,36^ (*FDR* < 0.1) (Supplementary Figure 4)^30^. In general, signal transduction pathways help coordinate the cellular response to external signals. Thus, the 35-gene set based on significant SNP interactions shares similar genes to glucocorticoid receptor blockage and common environmental response pathways, with links to cancer.

We sought to identify whether the interaction gene set was regulated by common transcription factors. Using transcription factor binding site information in EnrichR^30^ from ENCODE^37^ and ChEA^38^, we found that 10 of the 35 unique genes were targets of *NFE2L2* and its protein, NRF2 (*FDR* = 4.6 x 10^−4^) (**Figure 5E**). NRF2 mediates the cellular response to stress by transcriptionally activating an anti-oxidant and anti-inflammatory program^39,40^. However, NRF2 overexpression is linked to breast cancer development^41^ and tumorigenesis^42^ by continuing to provide protective benefit to cancerous cells. By analyzing GTEx samples^43^, we found that NRF2 gene expression (*NFE2L2*) is co-expressed with GR gene expression (*NR3C1*) across nearly all tissue types (*FDR* < 0.1). For breast tissue, co-expression is significantly stronger in female samples than in male samples (*r* = 0.59 versus r = 0.46; *P* = 0.028) (Supplementary Figure 5). By querying DSigDB^31^, we found that commonly-used corticosteroids, such as dexamethasone, hydrocortisone, or betamethasone, are associated with *NFE2L2* expression changes. Thus, glucocorticoid receptor modulation of NRF2 and germline alterations to the transcription factor targets of NRF2 may together harm cellular homeostasis and raise BC risk.

### Genotype stratification of breast cancer risk by corticosteroid use

Two SNPs reach *FDR* < 0.01 significance in their interaction with corticosteroids and BC risk. The SNP rs62119267, located in the *IGSF23* intron, has the most significant interaction with corticosteroid use with an interaction term *P*-value = 2.1 x 10^−8^, which is below the genome-wide significance threshold of *P* < 5 x 10^−8^ used routinely in GWAS. This SNP is associated marginally with BC risk in previous studies, as it is included within the PGS. However, it is far from the strongest signal: the marginal *P*-value = 0.004 in prior GWAS^25^. The second SNP, rs4784227 near the *TOX3* gene, is a very common variant in the population that has been strongly implicated previously in elevating BC risk.

Given the high significance of the rs62119267 interaction, we considered the effects of corticosteroids use on stratifying BC risk within carriers and non-carriers of rs62119267 variants. Within individuals who do not carry the minor allele, we found that there was no detectable difference in BC rates (**Figure 6A**). However, within rs62119267 carriers of at least one C allele, BC incidence for corticosteroid users was 18.2% during follow-up. In comparison, 5.1% of carriers who do not take corticosteroids develop BC (**Figure 6B**). As another statistic, the context-dependent per-allele effect within users is *HR* = 3.41 (95% CI: 2.16-5.42; *P* = 1.3 x 10^−6^), with no significant association in non-users. We observed no increase in false positive rate for the rs62119267-corticosteroid interaction when permuting disease status across samples. Furthermore, the rs62119267-corticosteroid interaction remains significant when matching non-users to corticosteroid users by propensity score (Supplementary Note 4). Overall, this large and robust interaction effect might explain some of the marginal GWAS effect estimate reported by the meta-analysis from Michailidou et al. (OR = 1.07; *P* = 0.004)^25^, with potential clinical implications for preventing cancer occurrence in women using corticosteroids and who carry the rs62119267 C allele.

**Figure 6:**
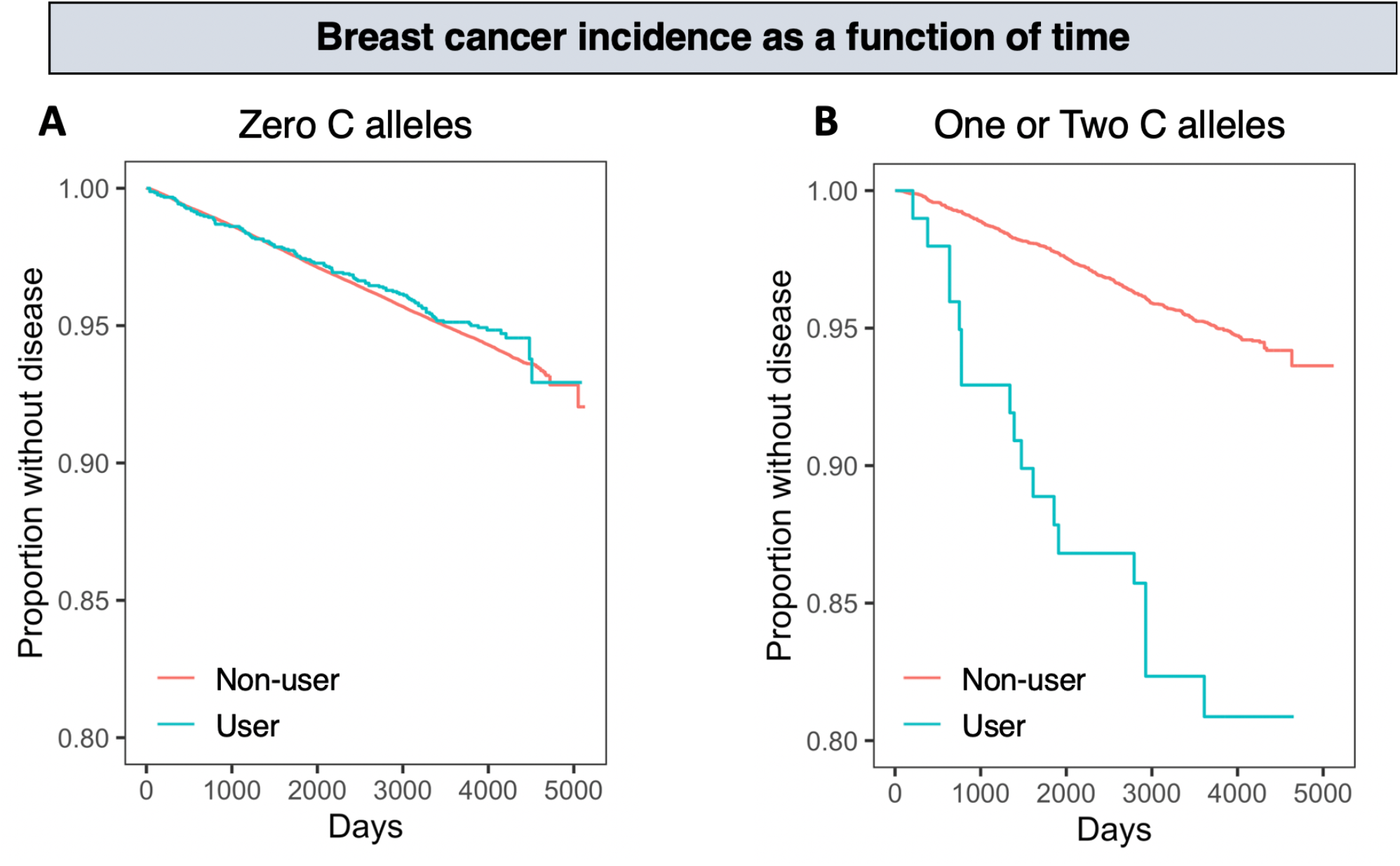
Risk stratification by rs62119267 genotype and corticosteroid use. In the left panel (A), we see individuals with no C allele at the rs62119267 genotype. In the right panel (B), we see carriers with at least one C allele. The proportion of individuals without breast cancer (Y-axis) is displayed as a function of time (X-axis). The red line indicates non-users of corticosteroids, while blue indicates the users of corticosteroids. In the carriers, we see that corticosteroid use leads to much higher incidence of breast cancer risk: nearly 20% of users developed breast cancer in UKB, compared to about 5% in non-users. No significant stratification was observed for women without C alleles.

While rs62119267 lies in the *IGSF23* intron, the nearest transcriptional start site is *PVR* (12,417 bp upstream). In an analysis of 27,738 whole blood samples, rs62119267 is significantly associated with *PVR* expression (nominal *P* = 2.4 x 10^−6^; *FDR* = 6.6 x 10^−3^), with some evidence of an expression association in a smaller cohort of 396 breast tissue samples from GTEx (*P* = 0.056). *PVR* encodes the immunoglobulin superfamily protein CD155, known as the poliovirus receptor. CD155 has an important role in helping cells evade the immune system^44,45^, which may be suppressed by corticosteroids, including facilitating the migration of immune cells through endothelial cells^46^. As well, CD155 is overexpressed in BC patients, and thus has been proposed as a potential immunotherapy target for cancer^47,48^. Together, our results highlight *PVR* as a potential context-dependent BC susceptibility gene.

## Discussion

Within UK Biobank, we performed a comprehensive gene-drug interaction analysis for breast cancer risk to discover corticosteroid use as a modulator of susceptibility loci across the genome. To date, cancer epidemiology studies of drug use and pharmacogenomic studies of particular drugs and their side effects have generally been performed separately. With the advent of UKB, a prospective cohort study pairing genetic, medication, and disease data for 500,000 individuals, it is now possible to apply genetic epidemiology approaches to study pharmacogenetic interactions across numerous germline polymorphisms, medications, and diseases. Thus, not only can we analyze how a person’s genetic profile interacts with their medication usage at-scale, we can also longitudinally explore why some users experience adverse outcomes or whether medications are available to reduce disease risk for people with genetic predispositions (**Figure 1**).

This opportunity does not come without its challenges. First and foremost, many cohorts such as UKB are not representative of randomized controlled trials (RCTs) for assessing the benefits and risks of drug use. As a result, interpreting the effects of a drug by comparing users versus non-users is not identical to performing a RCT, where users and non-users are assigned randomly (and non-users receive placebos) and the differences between the two groups are compared after time. As a result, in prospective cohorts, rapid identification of gene-drug interactions across the entire landscape of drugs and diseases in parallel is prone to confounding by indication, as the reason for taking a medication is unknown. Many common diseases, such as coronary artery disease or type 2 diabetes, have substantial co-morbidities and risk factors, and eliminating whether medication use or something else (which could also lead to medication use) affects risk will require careful considerations of confounders for each studied disease. For studying breast cancer, we attempt to mitigate potential issues that might arise by only studying drugs not known to be associated with breast cancer risk in the population (thus, any associations with drug use would be only found in genetic subtypes, which are randomized at birth) and performing propensity score analysis.

Second, the number of genetic variants to study can be enormous, the medication data available can be complex, and the interaction effects can be small and difficult to detect^5^. To overcome these challenges, we leveraged coordinated interactions, which aggregate antagonistic or synergistic interaction effects across SNPs (**Figure 3**). Sheppard et al.^24^ utilized this approach to find evidence for epistasis in phenotypes with no available evidence for individual pairwise SNP-SNP interactions, demonstrating increased statistical power. We too utilize this approach, along with a mapping of 3,603 distinct medication groups in UKB to a final 102 ATC groups, to perform a simple analysis of the interaction between genetics and medication usage (**Figure 2**).

In our study, we identify corticosteroids as a modulator of BC genetic risk (**Figure 5**). Corticosteroids are a class of steroid hormones with anti-inflammatory and immunosuppressive effects. Glucocorticoids, a dominant subset of corticosteroids, activate the glucocorticoid receptor, which has been shown to promote breast cancer metastasis^49^. However, it remains unclear whether genetically predisposed corticosteroid users have increased breast cancer development due to triggering cancer growth and proliferation or altering the immune infiltration within breast tissue^50^, or both. To functionally explore this interaction, we discovered SNPs in the PGS with individually strong corticosteroid interactions and mapped them to likely genes. Through a gene-set enrichment analysis, we found that this gene set overlapped with the gene signature of mifepristone, an approved glucocorticoid receptor antagonist currently used to terminate pregnancies, and signal transduction pathways relevant to environmental response and cancer. The anti-oxidant transcription factor NRF2 regulates many of our mapped interaction genes and is overexpressed in breast cancer cells^41^. NRF2 activates a transcriptional program to reduce reactive oxidative species and protect against DNA damage, and is a critical regulator of immunity^39,40,51^. This might suggest that gene-corticosteroid interactions affect a combination of cancerous proliferation and the immune system, although single-cell sequencing and functional experiments can provide a better window into this interplay.

There are several limitations with using the UKB dataset in our study. First, we are unable to test whether gene-corticosteroid interactions are associated with certain breast cancer subtypes, such as ER-positive versus ER-negative, since the information is not recorded within the UKB hospital record data. This could illuminate whether corticosteroids increase the risk of aggressive cancers or non-aggressive cancers in genetically susceptible individuals, and provide stronger characterization of phenotypic manifestations. Second, we do not have detailed information on medication usage for all individuals in UKB, which will allow an understanding of how dosage and duration affects risk. Third, other types of cancers are at much lower prevalence in UKB. While colorectal and prostate cancers are common in UKB men, no other cancer besides breast cancer is common in UKB women. For example, there are about 10% of the incident cases for ovarian cancer as compared to breast cancer in UKB. As a result, there is insufficient power to fully explore whether gene-corticosteroid interactions are associated with other cancers as well. However, present evidence suggests that other cancers have an at-best modest genetic correlation with BC^52^ and thus there will exist many BC-specific interactions.

Overall, we demonstrate that broad “coordinated” drug interactions with polygenic variation exist and are discoverable. Furthermore, we show that gene-drug interactions may have clinical relevance, such as the rs62119267 genotype stratifying breast cancer risk in corticosteroid users (**Figure 6**). These interactions are key to precision medicine ideas, where the objective is to best predict outcomes conditional on complex relationships between risk factors. Our methods provide a roadmap for future gene-drug studies in biobanks, as they can be extended across cancers, diseases, and cohorts for identifying gene-drug interactions; ultimately, in an effort to identify personalized recommendations of medication use.

## Methods

### UK Biobank data

The UK Biobank (UKB) team previously processed the UKB data^53^, and deposited for research in the scientific community. We accessed the UKB data under application ID 47137. We extracted the set of women with self-reported British European ancestry, and excluded all individuals with sex chromosome aneuploidy, excess heterozygosity, or outlier genotype missing rates. Breast cancer (BC) was diagnosed using procedural classifications (OPCS) and medical classifications (ICD9, ICD10) from longitudinal health record information, and self-reports during the baseline assessment. We used 174X in ICD9 information and C50X in ICD10 information. In OPCS data, we used B27, B28, or B29. We used self-report code 1002. We removed any individuals with a BC diagnosis prior to or reported at the baseline assessment. This left 212,335 women with no prior BC diagnosis at the time of baseline assessment, with 11,730 incident BC cases diagnoses after the baseline assessment (determined through the longitudinal health records) and 200,605 population controls. Individuals were censored at death, hospital records end date, or BC diagnosis. Imputed SNP data using were provided by the UKB team, calculated as described previously^53^, and were used for genetic analyses.

### Calculating BC polygenic scores

We use a polygenic score (PGS) to summarize breast cancer risk from genetic loci across the genome. The PGS can be computed using the standard calculation of a weighted sum of trait-associated SNPs (PGS = Σ_i_ *β*_i_ *X*_i_). We refer to *β*_i_ as the weight assigned to SNP *i*, and *X*_i_ as the number of minor alleles.

However, determining the appropriate weights for each SNP (*β*_i_) and which SNPs to include in the PGS leads to user and parameter bias, with variable accuracies as a result. As such, we use two breast cancer GWASs to calculate scores. Michailidou et al. released summary statistics from a GWAS meta-analysis of 61,282 cases across 68 cohorts. These were used to create original polygenic scores. Mavaddat et al. released PGS weights for two pre-computed scores in the PGS catalog based on a GWAS meta-analysis of 94,075 cases across 69 cohorts (an updated GWAS from Michailidou et al.). One score is based on hard-thresholding stepwise forward regression, while the other score is based on LASSO penalized regression. Neither score used UK Biobank to estimate or train SNP weights.

For both Michailidou et al. and Mavaddat et al. scores, we applied a quality control pipeline to handle ambiguous SNPs, account for potential sequencing errors, and focus on high-quality variants. We removed SNPs with missing chromosome, position, or effect size information. We removed multi-allelic SNPs, indels, SNPs with ambiguous strand flips (A/T, G/C), SNPs not present in UKB, and SNPs with poor imputation (INFO < 0.9). We flipped alleles if different strands were used in the summary statistics file compared to UKB (A/C in the summary statistics file, but T/G in UKB) and reversed alleles if necessary (A/C in file, C/A in UKB). For Michailidou et al. scores, odds ratio estimates provided by Michailidou et al. were transformed into *β* estimates via log transformation.

We intended to create multiple Michailidou et al. scores, and to select the optimal one for downstream analysis. To select SNPs for scoring 5 different Michailidou et al. scores, we used clumped summary statistics at 5 *P*-value thresholds: 0.05, 0.005, 0.0005, 10^−5^, 5 x 10^−8^. We set the r^2^ value = 0.1 and a window size equal to 250 kilobases. We performed clumping using a linkage disequilibrium panel of 503 European individuals from the 1000 Genomes project. In all, we created 7 scores: 5 Michailidou et al. scores and 2 Mavaddat et al. scores. All scores were standardized such that the mean was equal to zero and the standard deviation was equal to one.

To select the optimal Michailidou et al. and Mavaddat et al. scores, we fitted logistic regression models of breast cancer risk, calculated the Nagelkerke-R^2^ of each PGS, and selected the optimal Michailidou et al. score and optimal Mavaddat et al. score. We included body mass index, age, menopause status, number of births, at least one birth indicator, and the first 10 genotype principal components as covariates—we refer to these as the baseline covariates. We assessed the relationship between scores through pairwise Pearson’s correlation.

Due to modest correlation between scores, we combined the optimal Michailidou et al. score and the optimal Mavaddat et al. score into a combined score by taking the mean between the two. We use a Cox Proportional-Hazards model to calculate hazard ratios (*HR*) of PGSs. Individuals were binned into deciles, and disease incidence was calculated within each decile and compared to the 45^th^ – 55^th^ percentile individuals. The combined score was used in downstream analyses. Because of minimal competing risks, we use a Cox Proportional-Hazards model for all survival analyses as opposed to a Fine-Gray model.

### Identifying medication groups for analysis

At the baseline assessment, individuals were asked if they were regularly taking any medications. One limitation of this survey is that further information was not obtained; specifically, duration, dosage, and personal reasons for taking each medication are unknown.

We downloaded a mapping from UKB medications to ATC codes provided by Wu et al., and mapped medications to Level 4 ATC groups. We identified 106 ATC groups with at least 1000 users. We removed 4 ATC groups with a strong marginal association with BC risk (*FDR* < 0.01) by using 106 distinct Cox Proportional-Hazards models with the baseline covariates (minus PGS). Due to UKB not being a randomized controlled trial, these possibly include medications preferentially taken by individuals in high-risk groups which could be difficult to account-for in analysis and interpret within results.

### Testing for gene-drug interactions

We first tested for interactions between the breast cancer PGS and medication use on breast cancer risk. Using the 102 remaining ATC groups, we computed 102 Cox Proportional-Hazards models with baseline covariates, each medication group, and the combined PGS as main effects, with an interaction term between medication group and the combined PGS. We used false discovery rate for multiple hypothesis testing corrections. We fitted a logistic regression model with the same covariates to compute Nagelkerke-R^2^ as a measure of variation explained by the PGS within users versus non-users.

We extracted 3,342 SNP used to compute the combined PGS, and next tested for interactions between SNPs and corticosteroid use by using 3,342 Cox Proportional-Hazards models. We used the baseline covariates, one SNP, and S01BA use as main effects, and tested for the significance of the interaction term between the SNP and S01BA. We used false discovery rate for multiple testing corrections. To examine evidence for coordinated interactions, we assessed the concordance of the sign of the marginal GWAS effect estimates to the sign of the interaction effects in UKB at SNPs with an interaction *P*-value below 0.05. We calculated significance at risk-increasing and -decreasing SNPs by using a binomial test with probability equal to 0.5.

To further analyze the strongest SNP-corticosteroid interaction (rs62119267), we contrasted carriers of at least one C allele to non-carriers within survival plots and calculated per-allele odds ratio within corticosteroid users and non-users. We estimated false positive rates at this locus by permuting disease status, and calculating the false positive rate. Expression QTL data was obtained through the GTEx portal and the eQTLGen browser.

### Functional analysis of gene-corticosteroid interactions

We mapped SNPs with significant corticosteroid interactions to genes. If the SNP was located in the body of a protein-coding gene, the SNP was assigned to that gene. Otherwise, the SNP was mapped to the protein-coding gene with the highest variant-to-gene score in the Open Targets database. We uploaded the list of assigned genes to EnrichR for performing gene-set enrichment analyses. We assessed drug signature overlap by using the “DSigDB” database, gene ontology enrichments by using the “GO_Molecular_Function_2018” and the “GO_Biological_Process_2018” databases, and transcription factor regulation by using the “ENCODE_and_ChEA_Consensus_TFs_from_ChIP-X” database.

We used GTEx v8 data to test co-expression between GR and NRF2. We computed pairwise Pearson correlation between the GR gene expression (NR3C1) and NRF2 gene expression (NFE2L2) for 49 GTEx tissues with at least 70 samples. The 49 *P*-values across 49 tissue types were adjusted using false discovery rate. Finally, in breast tissue samples, we calculated correlation coefficients in males and females separately and compared the correlations by using one-sided test after a Fisher transformation.

## End Notes

## Data Availability

UK Biobank data was accessed under application number 47137. Computer code to reproduce the analyses is available at https://github.com/drewmard/druggene.

## Acknowledgements

We would like to thank Clark lab members, Elemento lab members, and Peter Kraft for helpful discussions surrounding this project. Figures were created with BioRender.com. Support was provided for A.R.M. by the NIH grant R01 ES029929.

## Author Contributions

A.R.M., A.G.C, and O.E. conceived and designed the study. A.R.M., S.K., and C.P. performed analyses. R.T. led key discussions. A.G.C. and O.E. supervised the study. All authors reviewed and approved the manuscript.

