## Supplementary Information for "A polygenic score-based approach to identify gene-drug interactions stratifying breast cancer risk"

**Marderstein et al.**

### Supplementary Figures

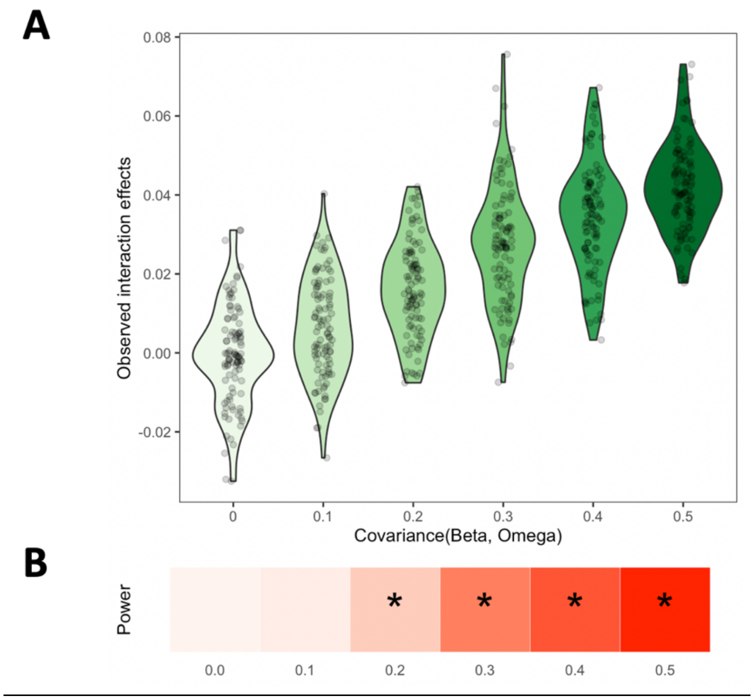


**Supplementary Figure 1: Detection of polygenic interactions are dependent on the coordination parameter.** (A) As the true covariance between main effects (Beta) and interaction effects (Omega) increase, the observed interaction effects in the coordinated interaction test increases. The x-axis displays Pearson correlation coefficients between the main and interaction effects. Each point is a new simulation. (B) A heatmap of the power to detect a significant interaction by using the coordinated interaction test. Darker red indicates higher power, where power is determined by calculating the proportion of simulations where $P < 0.05$. Asterisk indicates where the power was significantly different from 0.05 by a binomial test. As covariance increased, the power increased.

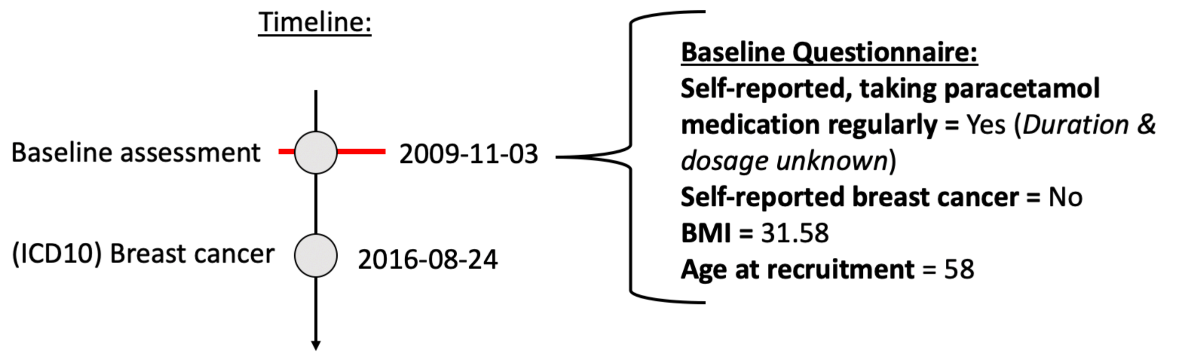


**Supplementary Figure 2: Visualization of the study data from UK Biobank.** For our study, we used an individual's data from the baseline questionnaire and longitudinal health records, in the form of ICD hospital codes.

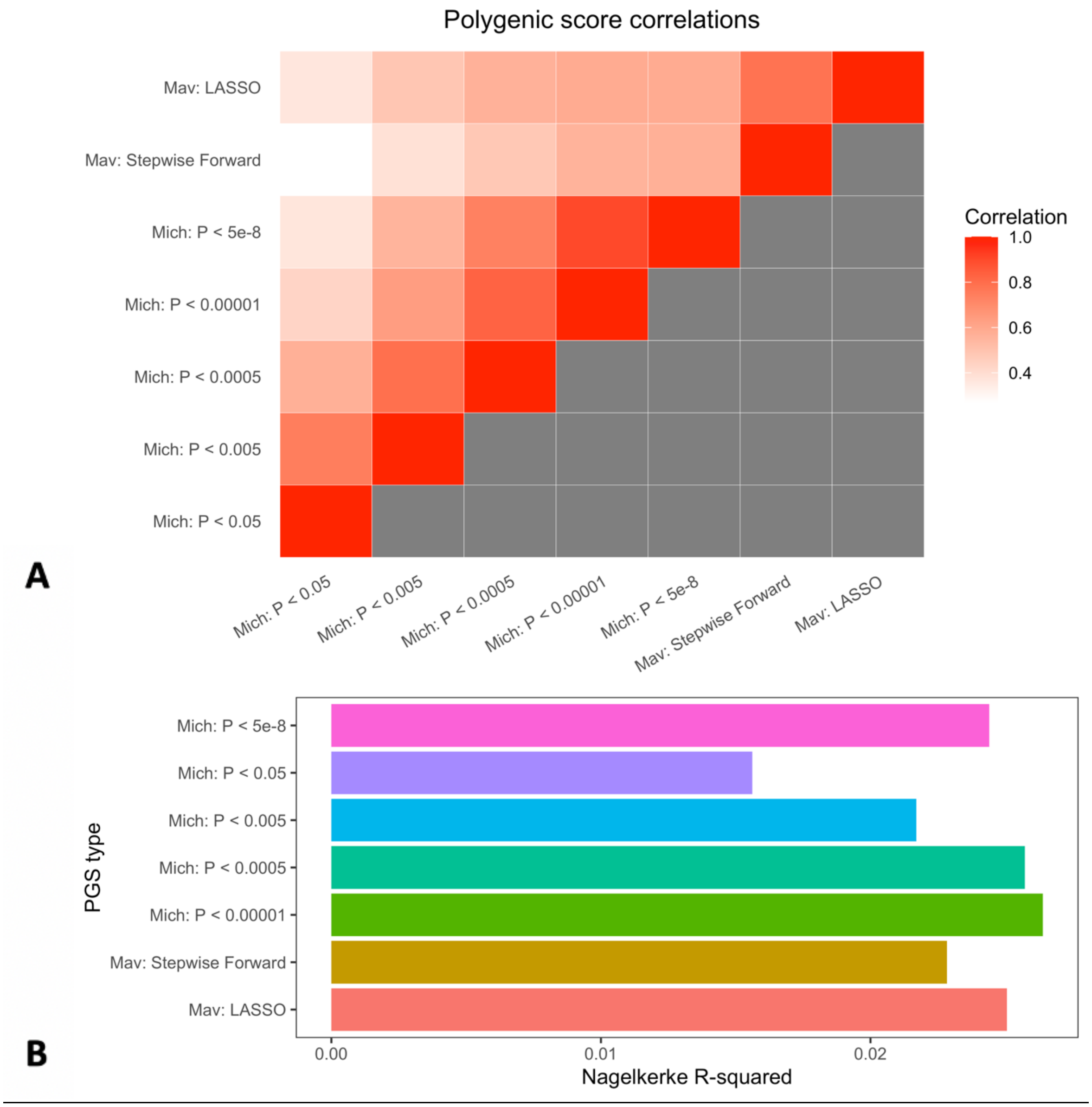


**Supplementary Figure 3: Correlation between scores and selecting the optimal predictor.** (A) Pairwise Pearson correlation between all Mich and Mav scores. (B) Nagelkerke R-squared of all scores, which were used to select the best Mich and Mav score.

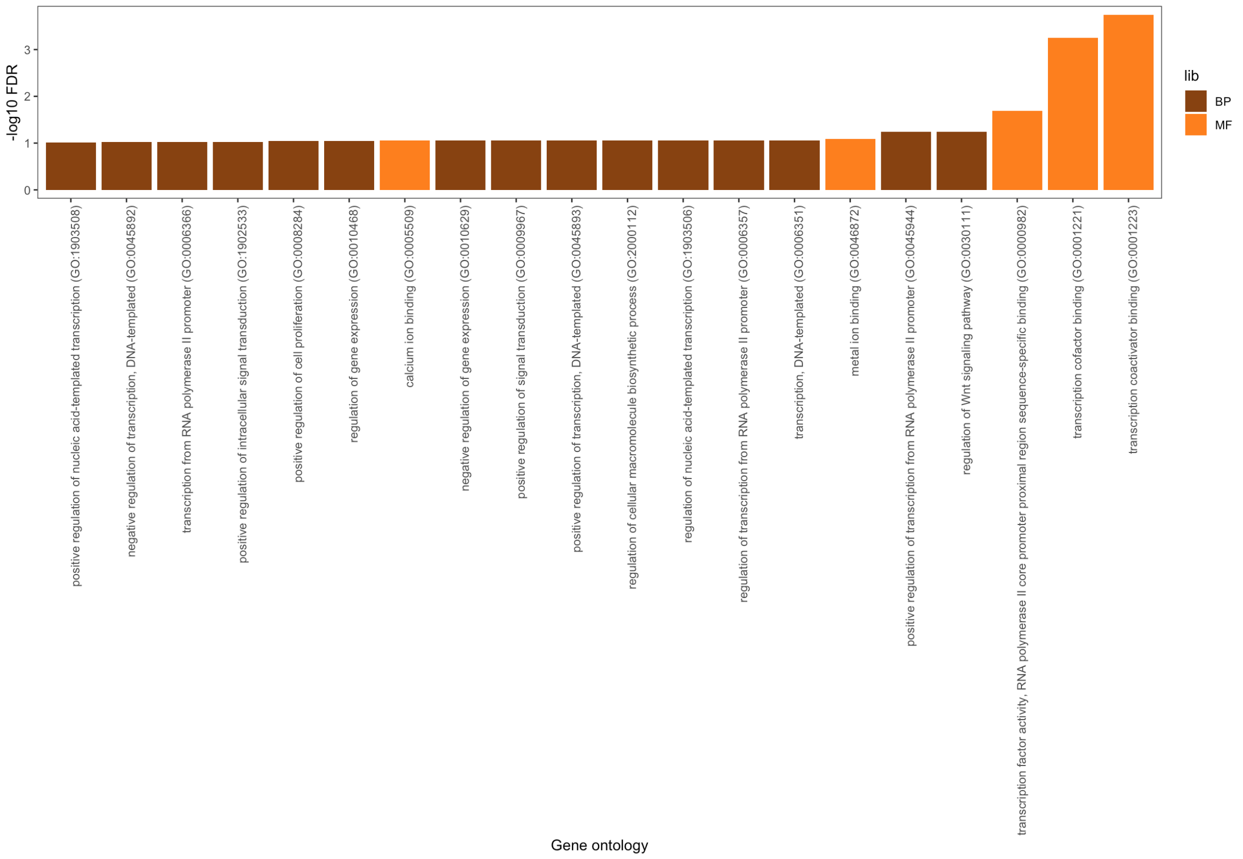


**Supplementary Figure 4: Gene ontology enrichment of genes implicated by SNP interactions.** Results based on an EnrichR analysis of the 35-interaction gene set for GO enrichment. Y-axis displays adjusted P-values via false discovery rate (FDR). MF = Molecular Function, BP = Biological Process.

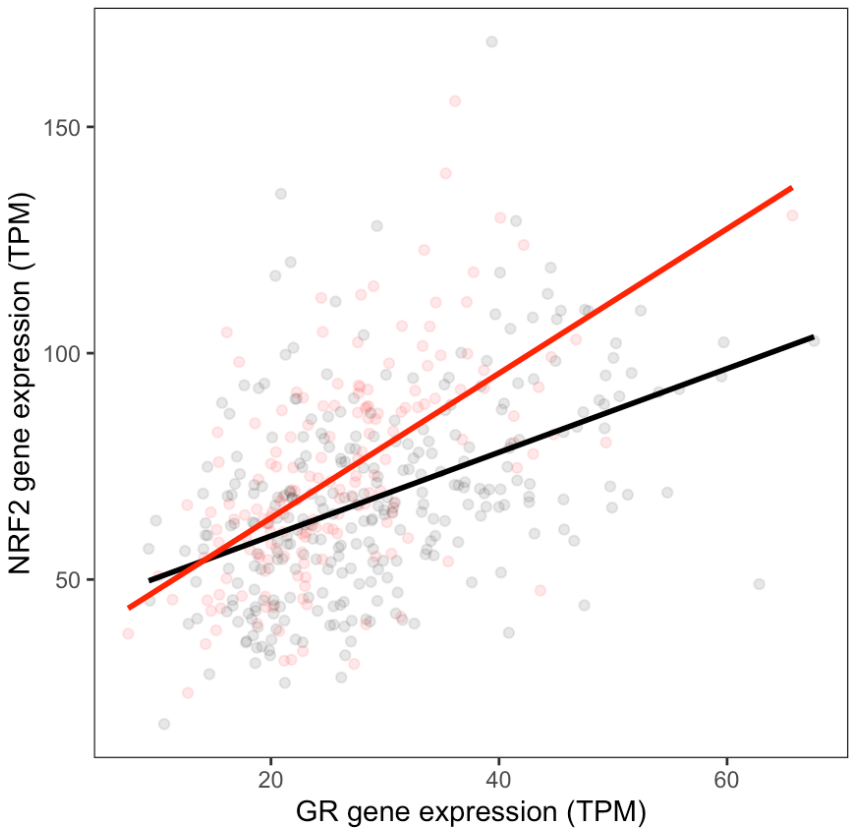


**Supplementary Figure 5: Co-expression of GR and NRF2 in breast GTEx samples by sex.** Scatterplot of *NR3C1* (x-axis) and *NFE2L2* expression (y-axis), colored by sex (black is for male samples, red is for female samples). Dark lines represent the slope of best fit within each sex.

### Supplementary Notes

#### Note 1: The coordinated interaction test by Sheppard et al.

Consider a pairwise polygenic interaction model between SNPs *X* and an environmental factor *Z* on a phenotype *y*:

$$y_i = \sum_{j=1}^{M} \beta_j X_{ij} + \sum_{j=1}^{M} \Omega_j X_{ij} Z_i \qquad \text{(Eq. 1)}$$

$\beta_j$ represents the main effect for SNP *j* on the phenotype *y*, $X_{ij}$ is the number of effect alleles for individual *i* at SNP *j*, $\Omega_j$ is the interaction effect between SNP *j* and the environmental factor *Z*, and $Z_i$ is the value of the environmental factor for individual *i*. To characterize the existence and impact of interactions on a phenotype across SNPs, existing methods often consider β and Ω as independent. We refer to these as uncoordinated interactions. However, Sheppard et al.[1] considers the covariance between β and Ω:

$$\gamma = \mathrm{Cov}(\beta,\Omega) \qquad \text{(Eq. 2)}$$

$\gamma$ describes the coordination parameter between main effects and interaction effects. When β and Ω are not independent, $\gamma \neq 0$. In practice, $\gamma$ can be estimated using the following regression model:

$$y \sim \alpha_1\, PGS + \alpha_2\, Z + \alpha_3\, PGS \times Z \qquad \text{(Eq. 3)}$$

PGS represents the polygenic score of additive main effects for the M SNPs, and Z is the environmental factor. The $\alpha_3$ estimate acts as a valid estimator for the coordination $\gamma$. Sheppard et al. provides theoretical proofs demonstrating the validity of this estimator, which also extends to the current gene-drug interaction work but is not presented within. We next describe simulations that assess the validity of a coordinated interaction test for detecting coordinated interactions, with no inflation under uncoordinated interactions.

**Note 2: Coordinated interaction tests are robust uncoordinated interactions**

Using simulations, we show how a polygenic-drug interaction test maintains false positive rates when the SNP interaction effects are uncorrelated with the SNP main effects.

In our simulations, we generate population genetic and disease status data. We generate 5,000 SNPs in 5,000 individuals, with minor allele frequencies randomly selected between 0.01 and 0.5. For SNP *j* with frequency $f_j$ ~ Uniform(0.01, 0.5), the genotype for individual *i* is $X_{ij}$ ~ Binom($f_j$, 2) (for simplicity, ignoring linkage disequilibrium). 10% of individuals in the population are users of medication *D*: $D_i$ ~ Binom(0.1,1). All 5,000 SNPs have non-zero additive effect on the underlying phenotypic liability: $\beta_j \sim N(0,1)$, with narrow-sense heritability $h^2 = 0.6$. 1,000 SNPs, representing 20% of all loci, have interaction effects with drug use to influence the phenotype, generated using $\Omega_j \sim N(0,1)$. To ensure that the first 1,000 values of β and Ω have no spurious covariance, we adjusted the Ω values to have correlation $r = 0$:

$$\Omega_{\text{new}} = \sigma_{\beta} \times \Omega' \qquad \text{(Eq. 4)}$$

where $\sigma_{\Omega}$ is the standard deviation of β, and $\Omega'$ is a vector of residuals after performing a linear model between Ω as the dependent variable and β as the independent variable. Interactions explain 10% of the phenotypic liability, while an independent environmental component (generated using a normal distribution) explains the remainder of the liability. Thus, the broad-sense heritability $H^2 = 0.7$. Phenotypic liability is calculated on a quantitative scale, containing the sum of scaled additive, epistatic, and environmental components.

Our simulation uses a multi-step process to assess the coordinated interaction test. A first population cohort is created. Within this cohort, a GWAS is performed using linear regression to estimate the marginal association between a SNP and the phenotype. Second, a new population cohort is created using the same parameter values. Within this new cohort, a polygenic score for all individuals is calculated by using the weights from the GWAS summary statistics in the first cohort. Finally, the interaction between polygenic score and medication use is tested for by including an interaction term within a linear regression model applied to the second cohort. The null hypothesis is rejected when $P < 0.05$ for the interaction term. 100 iterations are performed, and the null hypothesis rejection rate (proportion of iterations where $P < 0.05$) is used as the false positive rate (Supplementary Figure 1).

Across the 100 iterations, we found no evidence for increased test rejection rate. In all, only 6% of iterations led to test rejection compared to the expected 5% (Kolmogorov-Smirnov test for skewed *P*-value distribution: $P > 0.05$). Hence, a coordinated interaction test is robust to the presence of widespread uncoordinated interactions.

**Note 3: Correlated main and interaction effects power coordinated interaction test**

Next, we considered a simulation setting where main and interaction effects are correlated. The same simulation was performed, except the 1000 values of Ω were generated to have a defined correlation to the first 1000 values of β. This was performed using the following:

$$\Omega_{\text{new}} = r \times \beta \times \sigma_{\Omega'} + \sqrt{1 - r^2}\, \sigma_{\beta} \times \Omega' \qquad \text{(Eq. 5)}$$

$\sigma_{\Omega}$ is the standard deviation of β, $\Omega'$ is a vector of residuals after performing a linear model between Ω as the dependent variable and β as the independent variable, and *r* is the pre-defined value such that the first 1,000 values of $\Omega_{\text{new}}$ and β have a Pearson correlation coefficient equal to *r*. Furthermore, in these simulations, the null hypothesis rejection rate across iterations is referred to as the power, since there is now non-zero covariance between Ω and β. The variance explained by the interactions is identical to the previous uncoordinated interaction simulations.

We tested $r = [0, 0.1, 0.2, 0.3, 0.4, 0.5]$. We find that, as $r$ increases, the estimated interaction effects increase on average; in parallel, the power increases as well (Supplementary Figure 1). Since the interaction term showed no enrichment for statistical significance when $r = 0$ under the same narrow-sense and broad-sense heritability values (Supplementary Note 2), we can conclude that correlations between main and interaction effects drives the power to detect polygenic-drug interactions.

**Note 4: Propensity score matching for analyzing corticosteroid effects**

We explored whether underlying factors and indications that drive why individuals take corticosteroids confound the BC risk effect of corticosteroid users versus non-users. We considered asthma, allergy, Crohn's disease, ulcerative colitis, rheumatoid arthritis, systemic lupus erythematosus diagnosis, Townsend deprivation index, C-reactive protein serum measurement, and the baseline covariates as potential factors that lead to corticosteroid use and also be associated with BC risk. We matched non-users to the corticosteroid users using a nearest neighbor generalized linear model approach implemented in the MatchIt R package[2]. We matched non-users to users such that there are equal numbers of both groups (1:1 ratio). After matching on propensity score of corticosteroids, we performed three BC interaction tests, between: (1) polygenic score and corticosteroid use, (2) rs62119267 (*PVR*) and corticosteroid use, and (3) rs4784227 (*TOX3*) and corticosteroid use. We performed a Cox-proportional hazards model for the interaction test with baseline covariates. We calculated *P*-values by repeating this entire procedure across 100 iterations and computing the proportion of iterations where the interaction test was antagonistic ($HR < 1$). Overall, we found that all three interaction tests remained statistically significant in the propensity score matching analysis: the PGS-corticosteroid interaction had $P = 0.02$, with either SNP interaction present across all iterations ($HR > 1$ in all tests; $P < 0.01$).
